# A Comparative Study of Target Reconstruction of Ultra-High-Resolution CT for Patients with Corona-Virus Disease 2019 (COVID-19)

**DOI:** 10.1101/2020.06.04.20119206

**Authors:** Shao-mao Lv, Yu Lin, Jiang-he Kang, Shao-yin Duan, Wei-guo Zhang, Jin-an Wang

**Affiliations:** Department of Radiology, Zhongshan Hospital, Xiamen University, Xiamen 361004, China; Department of Radiology, Daping Hospital, Army Medical University, Chongqing 400010, China

**Keywords:** U-HRCT, 1024 matrix, Target Reconstruction, COVID-19

## Abstract

**Background:** The corona-virus disease 2019 (COVID-19) pandemic has caused a serious public health risk. Compared with conventional high-resolution CT (C-HRCT, matrix 512), ultra-high resolution CT (U-HRCT, matrix 1024) can increase the effective pixel per unit volume by about 4 times. Our study is to evaluate the value of target reconstruction of U-HRCT in the accurate diagnosis of COVID-19.

**Methods:** A total of 13 COVID-19 cases, 44 cases of other pneumonias, and 6 cases of ground-glass nodules were retrospectively analyzed. The data were categorized into groups A (C-HRCT) and B (U-HRCT), following which iDose^4^-3 and iDose^4^-5 were used for target reconstruction, respectively. CT value, noise, and signal-to-noise ratio (SNR) in different reconstructed images were measured. Two senior imaging doctors scored the image quality and the structure of the lesions on a 5-point scale. Chi-square test, variance analysis, and binarylogistic regression analysis were used for statistical analysis.

**Results:** **U-**HRCT image can reduce noise and improve SNR with an increase of the iterative reconstruction level. The SNR of U-HRCT image was lower than that of the C-HRCT image of the same iDose^4^level, and the noise of U-HRCT was higher than that of C-HRCT image; the difference was statistically significant (*P*< 0.05). Logistic regression analysis showed thatperipleural distribution, thickening of blood vessels and interlobular septum, and crazy-paving pattern were independent indictors of the COVID-19 on U-HRCT. U-HRCT was superior to C-HRCT in showing the blood vessels, bronchial wall, and interlobular septum in the ground-glass opacities; the difference was statistically significant (P < 0.05).

**Conclusions:** Peripleural distribution, thickening of blood vessels and interlobular septum, and crazy-paving pattern on U-HRCT are favorable signs for COVID-19. U-HRCT is superior to C-HRCT in displaying the blood vessels, bronchial walls, and interlobular septum for evaluating COVID-19.

## Background

The outbreak of the novel corona-virus disease 2019 (COVID-19) caused by severe acute respiratory syndrome coronavirus 2 (SARS-CoV-2) has spread worldwide[1]. The COVID-19 pandemic has caused a serious public health risk. COVID-19 is characterized by high infectivity and a typical clinical features[2]. The diagnosis of COVID-19 relied on virus nucleic acid detection with a high false negative rate [3].

High resolution computed tomography (HRCT) is a convenient and accessible method for early screening and diagnosis of viral pneumonia [4]. Typical chest HRCT findings for COVID-19 included the ground glass lesions, enlarged blood vessels and “crazy paving signs” [5].

Compared with conventional high-resolution CT (C-HRCT, with matrix of 512×512), ultra-high resolution CT (U-HRCT, with matrix of 1024×1024) can increase the effective pixel per unit volume by about 4 times in the same field of view (FOV) [6]. The UHRCT can not only reveal each ground glass lesion with sub-millimeter precision, but also quantify the extent and severity of each lesion[7,8].

Thus, the purpose of this study was to evaluate the value of U-HRCT in the quantitative assessment of COVID-19 and to compare the radiological patterns of U-HRCT and C-HRCT.

## Methods

### Patients

The study conducted a retrospective analysis of 63 suspected COVID-19 patients examined using CT at our hospital between January 22 and February 15, 2020. The patients included 35 males and 28 females aged between 6–69 years, with an average age of (35 ± 10.6) years. Among them, 13 cases were confirmed as COVID-19, 44 cases were diagnosed with other pneumonias, and 6 cases were diagnosed with ground-glass opacities.

### CT scans

The Ingenuity CT scanner (Philips Healthcare, the Netherlands) was used. The patient was placed in a supine position with breath hold during the scan. The scanning range was from the apex to the base of the lung. Scanning parameters included the following: tube voltage120 kV; mAs: DoseRight Index 15–20; collimation64 × 0.625 mm; pitch, 1.2; slice thickness 2 mm; interval 2 mm; FOV 35 cm. The raw data acquired were categorized into groups A (512 matrix) and B (1024 matrix). Each group was reconstructed using iDose^4^-3 and iDose^4^-5 iterative reconstructions within the same small FOV with a slice thickness and interval of 1 mm.

### Image analysis

#### Analysis of typical signs

A group of images was selected at random, and the CT data of all patients were evaluated by two radiologists with over 10 years of experience while blinded to each other’s results. The assessment of lesions included the following: (1) distribution (distance from the pleura); (2) number(single or multiple); (3) density (pure ground-glass opacities, mixed ground-glass opacities, and consolidation); and (4) Internal structure (air bronchograms sign, vascular thickening, interlobular septal thickening, and crazy-paving patterns in the lesions).

#### Objective scoring of image quality

Each group of images was transferred to the Philips IntelliSpace Portal workstation. The relevant information was locked, and the iDose3 and iDose5 groups of images were reconstructed, measured, and evaluated in the fixed lung window (window width 1600 HU, window level −600 HU). A region of interest (ROI) (50 mm^2^ approximate area) in the lung tissue at the level of the tracheal carina and left atrium was selected (avoiding lung markings and lesion areas), the standard deviation (SD) was measured, and the average CT value was recorded to calculate the signal-to-noise ratio (SNR). Each ROI was measured three times, and the average value was recorded. SNR = CTn/SDn, where CTn and SDn are the average CT and SD values of the lung tissue, respectively. The SD value is the objective noise of the image.

#### Subjective scoring of image quality

The two radiologists, with over 10 years of work experience, quantitatively evaluated image quality under the same magnification ratio. Differences in opinion were resolved by consensus. Image quality was scored on a 5-point scale. The evaluation included image noise, presentation of the bronchovascular bundles 2 cm from the pleura, presentation of the lesion and its internal structure, and feasibility of diagnosis. For scoring, 5 points: clear lesion and vascular structure with no artifacts; 4 points: acceptable lesion and vascular structure with a small number of artifacts; 3 points: unclear lesion and vascular structure with more artifacts, but diagnosis was unaffected; 2 points: unclear lesion and vascular structure with significant artifacts making diagnosis unsatisfactory; 1 point: unclear lesion and vascular structure, and multiple types of artifacts making diagnosis unsatisfactory.

#### Statistical analysis

The SPSS 22.0 software was used for analysis. Measurement data were expressed as 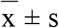. ANOVA was performed on the SD and SNR values of each group of images. Differences with *p*< 0.05 were considered statistically significant. Pairwise comparisons between the groups were performed. Count data were expressed as number of cases and percentage, and the Pearson χ^2^ test was used for comparisons between groups. Binary logistic regression was used for analysis of independent risk factors of CT diagnosis of COVID-19. The receiver operating characteristic (ROC) curve was used for analysis of the sensitivity and specificity of risk factors of COVID-19 prediction. Medcalc software (version 17.6) was used for comparison and plotting the efficacy of the area under the curve (AUC). Differences with *p*< 0.05 were considered statistically significant.

## Results

### Clinical characteristics

A total of 63 cases were included in the study. Some patients underwent CT re-examinations, and a total of 75 groups of CT data were obtained for analysis. The 13 confirmed cases of COVID-19 were primarily imported cases or had a history of contact (12/13). There were 59 cases of other types of pneumonias and 6 cases of ground-glass nodules.

### CT presentation

Analysis of the CT presentation of the lesions included the distribution and density of lesions and the condition within the lesions (Table 1). With respect to lesion distribution, COVID-19 lesions were mainly distributed within 2 cm of the pleura (87.5%), whereas lesion distribution was non-specific in other pneumonias. COVID-19 lesions were mainly pure (75%) or mixed (18.75%) ground-glass opacities, whereas lesions from other pneumonias were mainly of mixed density (64.41%). Air bronchograms, vascular thickening, interlobular septal thickening, and crazy-paving signs were present in COVID-19 lesions, at frequencies of 81.5%,87.5%, 87.5%, and 93.75%, respectively. Air bronchograms were also common in non-COVID-19 pneumonias (57.63%). The differences in all radiologicalsigns, except the number of lesions, were statistically significant between COVID-19 and non-COVID-19 lesions (*p*< 0.05).

**Table 1.**
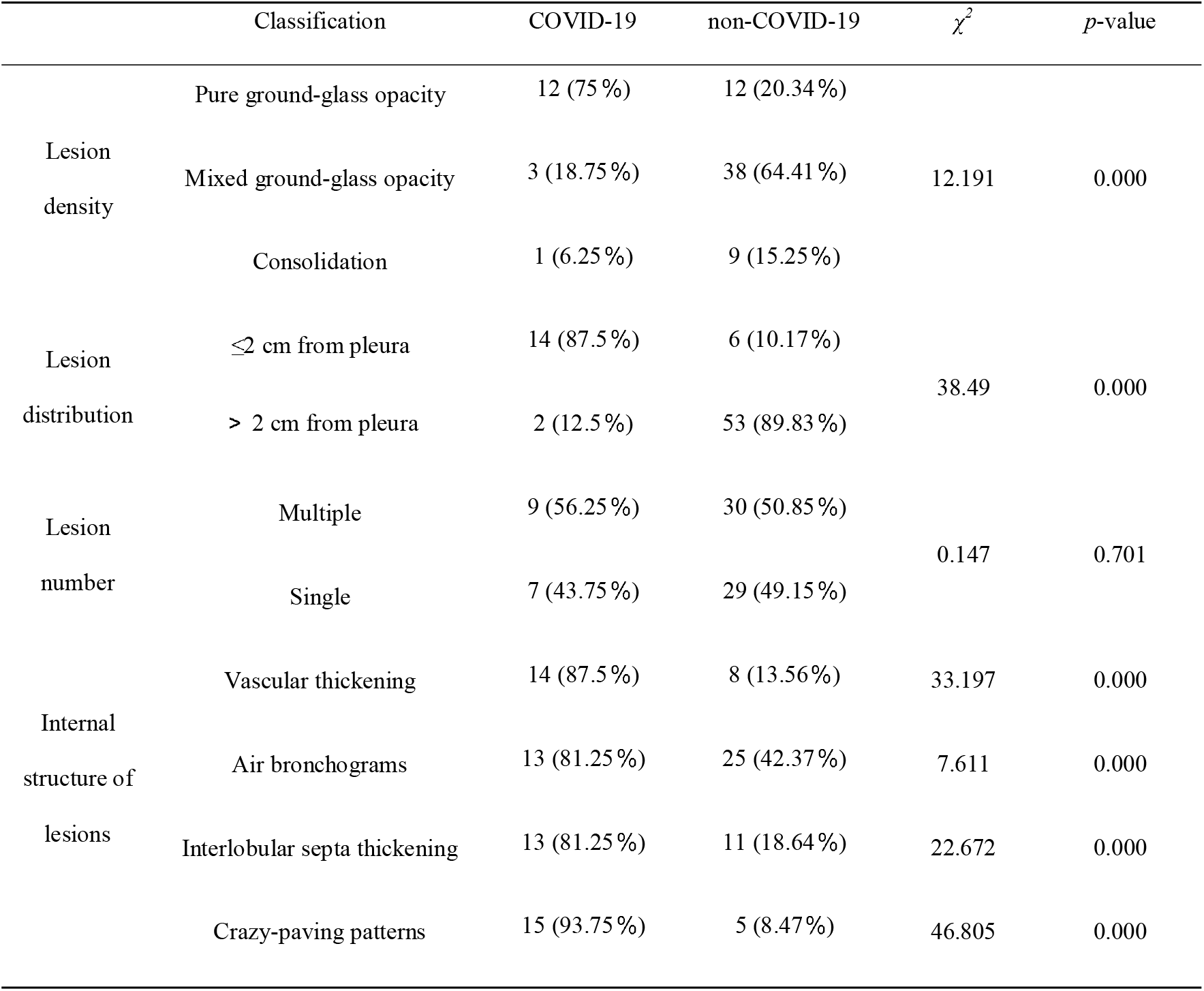
Analysis of CT signs of COVID-19 and non-COVID-19 lesions

| | Classification | COVID-19 | non-COVID-19 | $\chi^2$ | *p*-value |
| --- | --- | --- | --- | --- | --- |
| Lesion density | Pure ground-glass opacity | 12 (75 %) | 12 (20.34 %) | 12.191 | 0.000 |
|  | Mixed ground-glass opacity | 3 (18.75 %) | 38 (64.41 %) |  |  |
|  | Consolidation | 1 (6.25 %) | 9 (15.25 %) |  |  |
| Lesion distribution | ≤2 cm from pleura | 14 (87.5 %) | 6 (10.17 %) | 38.49 | 0.000 |
|  | > 2 cm from pleura | 2 (12.5 %) | 53 (89.83 %) |  |  |
| Lesion number | Multiple | 9 (56.25 %) | 30 (50.85 %) | 0.147 | 0.701 |
|  | Single | 7 (43.75 %) | 29 (49.15 %) |  |  |
| Internal structure of lesions | Vascular thickening | 14 (87.5 %) | 8 (13.56 %) | 33.197 | 0.000 |
|  | Air bronchograms | 13 (81.25 %) | 25 (42.37 %) | 7.611 | 0.000 |
|  | Interlobular septa thickening | 13 (81.25 %) | 11 (18.64 %) | 22.672 | 0.000 |
|  | Crazy-paving patterns | 15 (93.75 %) | 5 (8.47 %) | 46.805 | 0.000 |

### Logistic regression analysis

Lesion distribution and density, and structural features within the lesion were entered into a logistic regression analysis model to evaluate the independent risk factors of CT diagnosis of COVID-19. The analysis showed that lesion distribution, vascular thickening, interlobular septal thickening, and crazy-paving patterns had value for COVID-19 diagnosis (OR = 0.001, *p* = 0.003; OR = 43.212, *p* = 0.008; OR = 25.962, *p* = 0.022; OR = 258.081, *p* = 0.0001, respectively). However, lesion density and air bronchograms had minimal predictive value, and there was no statistically significant difference between two groups (Table 2).

**Table 2.** Logistic regression analysis of CT signs of COVID-19

| Indicator | B-value | OR (95%CI) | *p*-value |
| --- | --- | --- | --- |
| Lesion distribution | -4.508 | 0.011 (0.0006,0.216) | **0.003** |
| Lesion density | -1.114 | 0.3289 (0.054,1.990) | 0.226 |
| Vascular thickening | 3.766 | 43.213 (2.677,697.430) | **0.008** |
| Air bronchograms | 2.235 | 9.342 (0.675,129.208) | 0.095 |
| Interlobular septa thickening | 3.257 | 25.9615 (1.589,424.151) | **0.022** |
| Crazy-paving pattern | 5.553 | 258.081 (16.502,4036.160) | **0.000** |

**Table 3.**
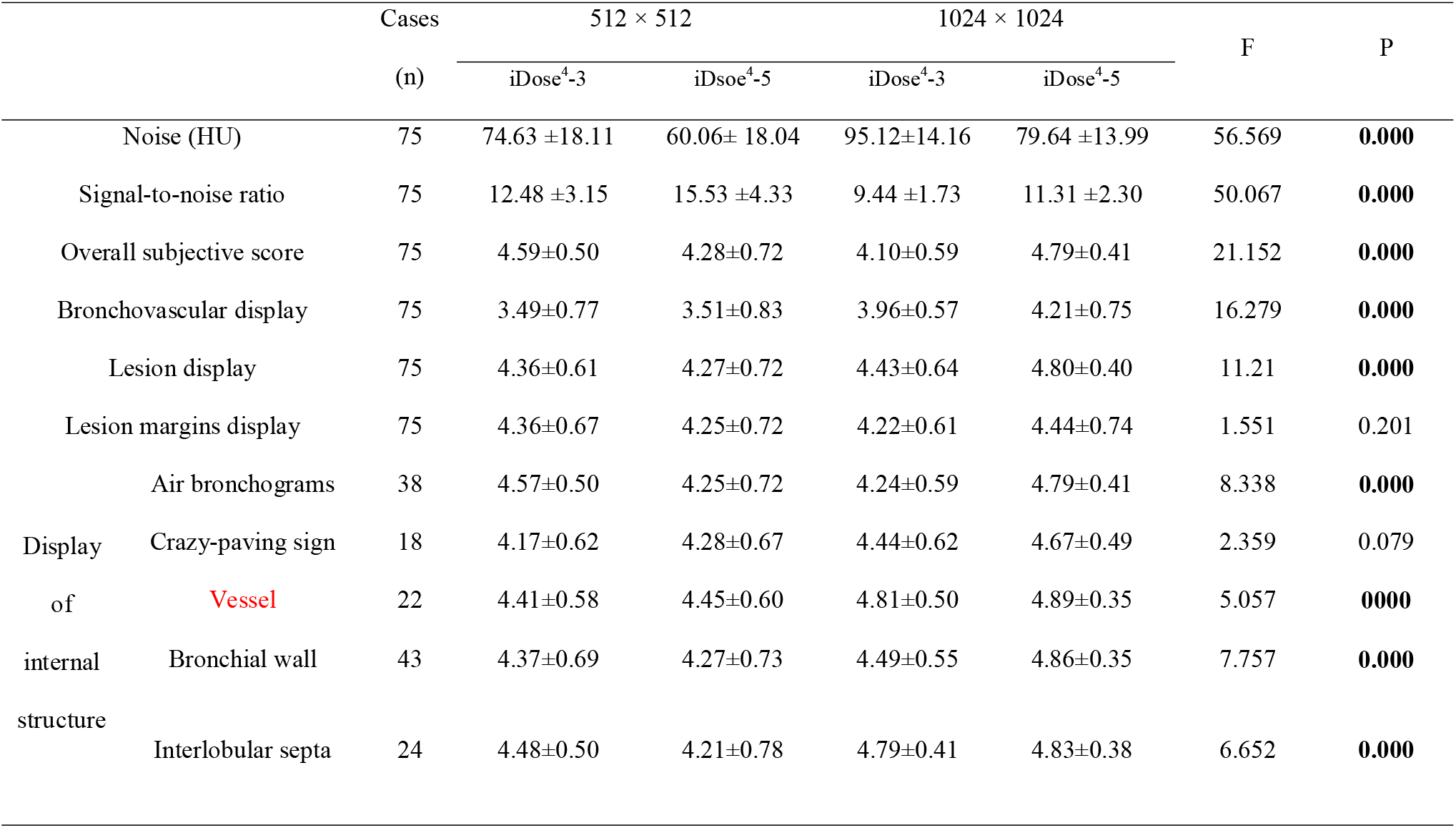
C-HRCT and U-HRCT image quality evaluation

### ROC analysis

ROC curve analysis showed that the AUC of lesion distribution, vascular thickening, interlobular septal thickening, and crazy-paving patterns were 0.887, 0.870, 0.813, and 0.926, respectively, with sensitivities / specificities of 84.8% / 77.9%, 90.1% / 81.4%, 85.1% / 92.5% and85.4% / 91.5% (Figure 1).

**Fig. 1.**
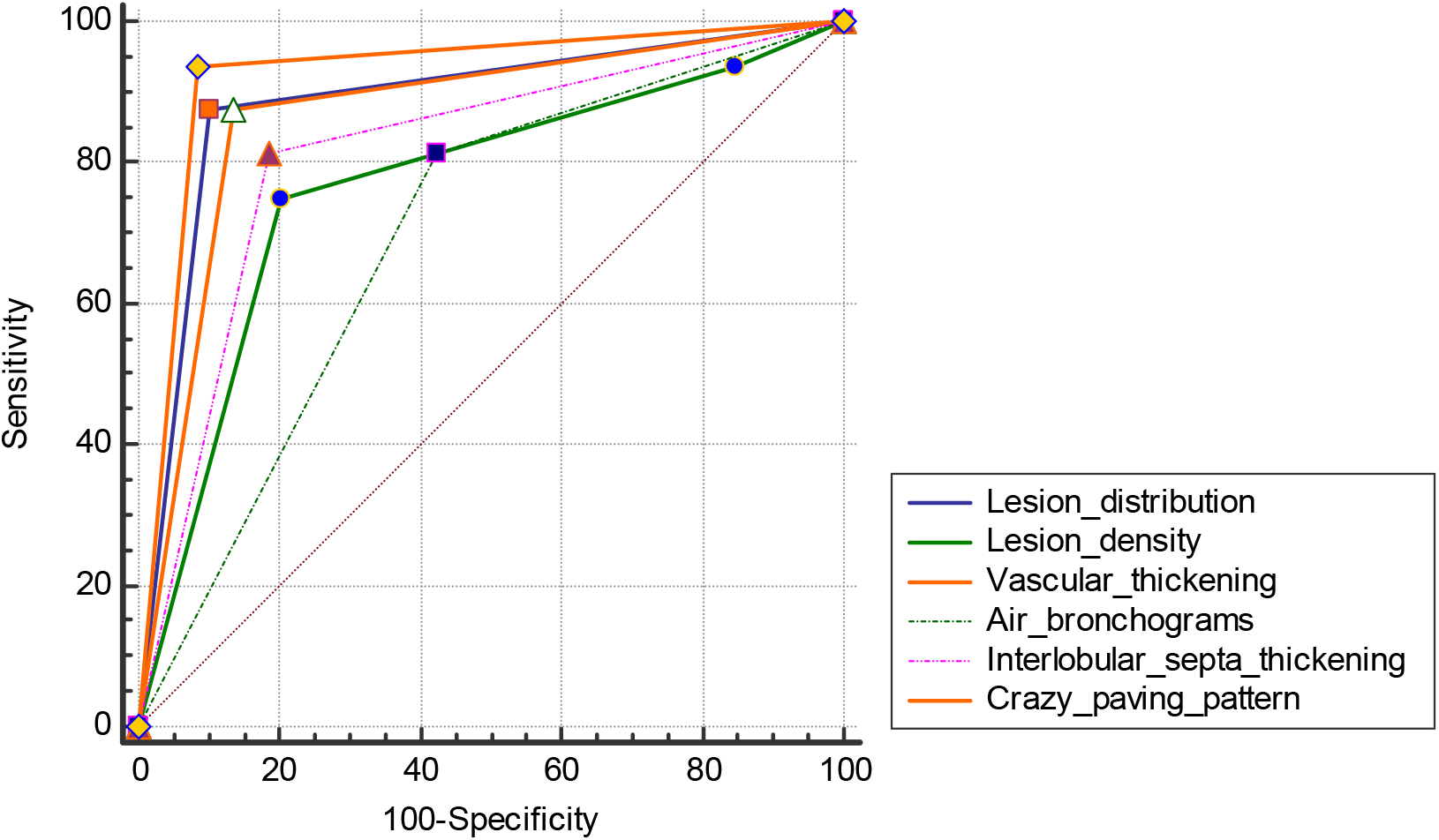
ROC curve of lesion distribution,density, and internal structure.

### C-HRCT and U-HRCT image quality scoring

With respect to objective image quality scores, the SD in U-HRCT was greater than that of C-HRCT, and the SNR was lower than that of C-HRCT; the difference was statistically significant (*p*< 0.05) (Table 2). With increased iteration levels, SD decreased, and SNR increased. At the same iteration level, the SD of U-HRCT was greater than that of C-HRCT, and the SNR was lower than that of C-HRCT. Pairwise comparisons showed that there was no statistically significant difference in image quality between U-HRCT using iDose5 iteration and C-HRCT using iDose3 conditions (*p* > 0.05).

The iDose^4^-5 iteration had the highest U-HRCT score for displaying the lesion structure and vascular structure of lung tissue. Except for display of lesion margins and crazy-paving patterns within the lesion, the differences in subjective scores were statistically significant (*p* < 0.05). Pairwise comparisons showed that the C-HRCT scores at high iteration levels were lower than those at low iteration levels for display of the internal structure of some lesions; there was no statistically significant difference between the two iteration levels (*p* > 0.05). There was no statistically significant difference in subjective scores for display of air bronchograms between U-HRCT and C-HRCT (*p*> 0.05).

## Discussion

COVID-19 is highly infectious and progresses rapidly. In severe cases, acute respiratory distress syndrome, respiratory failure, and even death may occur[9]. Diagnosis depends primarily on comprehensive consideration of epidemiology, clinical manifestations, and imaging and laboratory data. Chest CT, especially HRCT, is valuable in the diagnosis of suspected cases of COVID-19[10,11]. Although the imaging presentation of COVID-19 is similar to that of other viral pneumonias, the differential diagnosis is more difficult.However, COVID-19 could manifest some characteristic imaging signs, especially when ground-glass opacities werepresent[12,13].

In the confirmed COVID-19 cases in the present study, the lesions had a primarily subpleural distribution and had a ground-glass appearance. Vascular thickening, air bronchograms, interlobular septal thickening, and crazy-paving patterns were common in COVID-19 lesions. Air bronchograms were also common in other pneumonias, and display of the bronchial walls is vital in the diagonise. The results of small-sample binary logistic regression showed that the distribution of ground-glass lesions, vascular thickening, interlobular septal thickening, and crazy-paving patterns have some value for diagnosing COVID-19. COVID-19 could be more accurately diagnosed when taken together with the patient’s epidemiological and radiological characteristics, which is generally consistent with the findings of previous studies[14].

Display of the internal structure of ground-glass lesions is highly valuable for the diagnosis of COVID-19[15]. In practice, display of the internal details of these lesions is associated with the scanning technology used. U-HRCT uses a 1024 × 1024 matrix, allowing it to better display the morphological characteristics of pulmonary lesions[6,7,8]. Therefore, 1024 matrix U-HRCT images were retrospectively reconstructed and compared with C-HRCT in order to more accurately display the internal structure of ground-glass lesions and improve the accuracy of COVID-19 diagnosis. Compared to C-HRCT, U-HRCT has increased SD and decreased SNRin the present study.

In addition, iterative reconstruction techniques can also reduce SD and improve SNR[16-17].However, the reconstruction speed will be slower with higheriDose^4^iteration level, and the densities of the lesion and normal tissues tend to become homogeneous, which affects the observation of the internal structure of the lesion[18].Therefore, selection of an appropriate iteration level is essential.

In the present study, the image quality of U-HRCT and C-HRCT with the iDose^4^-3 and iDose^4^-5 iterative levels were compared and analyzed. We believe that the use of iDose^4^-5 iterative reconstruction can reduce noise and improve image quality, as well as improve spatial and density resolution of images. Additionally, the reconstruction speed of iDose^4^-5 is suitable for clinical applications. The iDose^4^-5 iteration level has the highest U-HRCT score for displaying the lesion and vascular structures of the lung tissue; it is particularly suited for vascular thickening, interlobular septal thickening, and other signs in the internal structure of the lesion that are important for diagnosis (Figures 2–4).

**Fig. 2.**
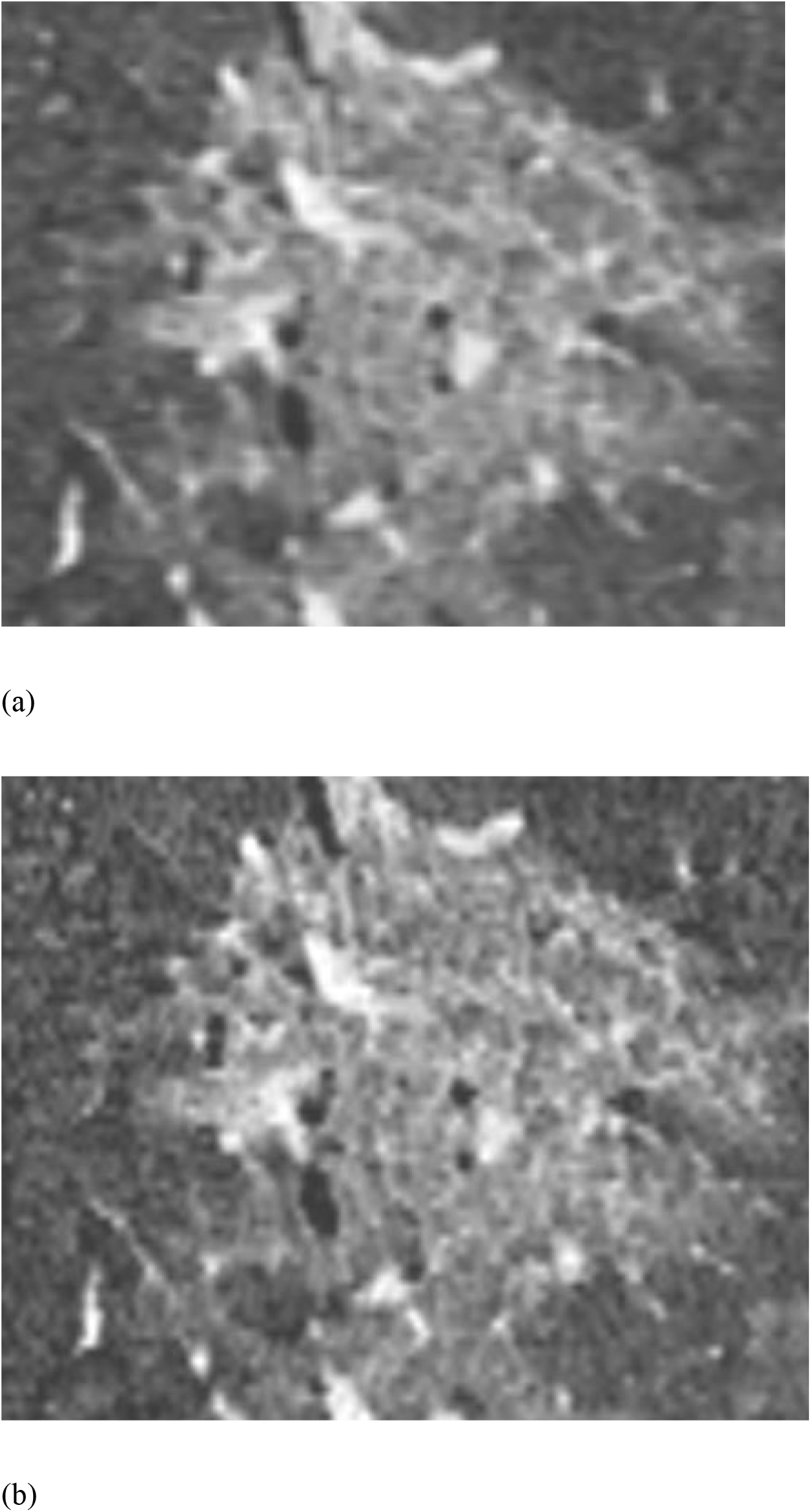
Coronal reconstruction of (a) C-HRCT and(b) U-HRCT images of a female 43-year-old patient with COVID-19.Bronchial walls and crazy-paving patterns are clearly displayed inU-HRCT but not clearly displayed inC-HRCT

**Fig. 3.**
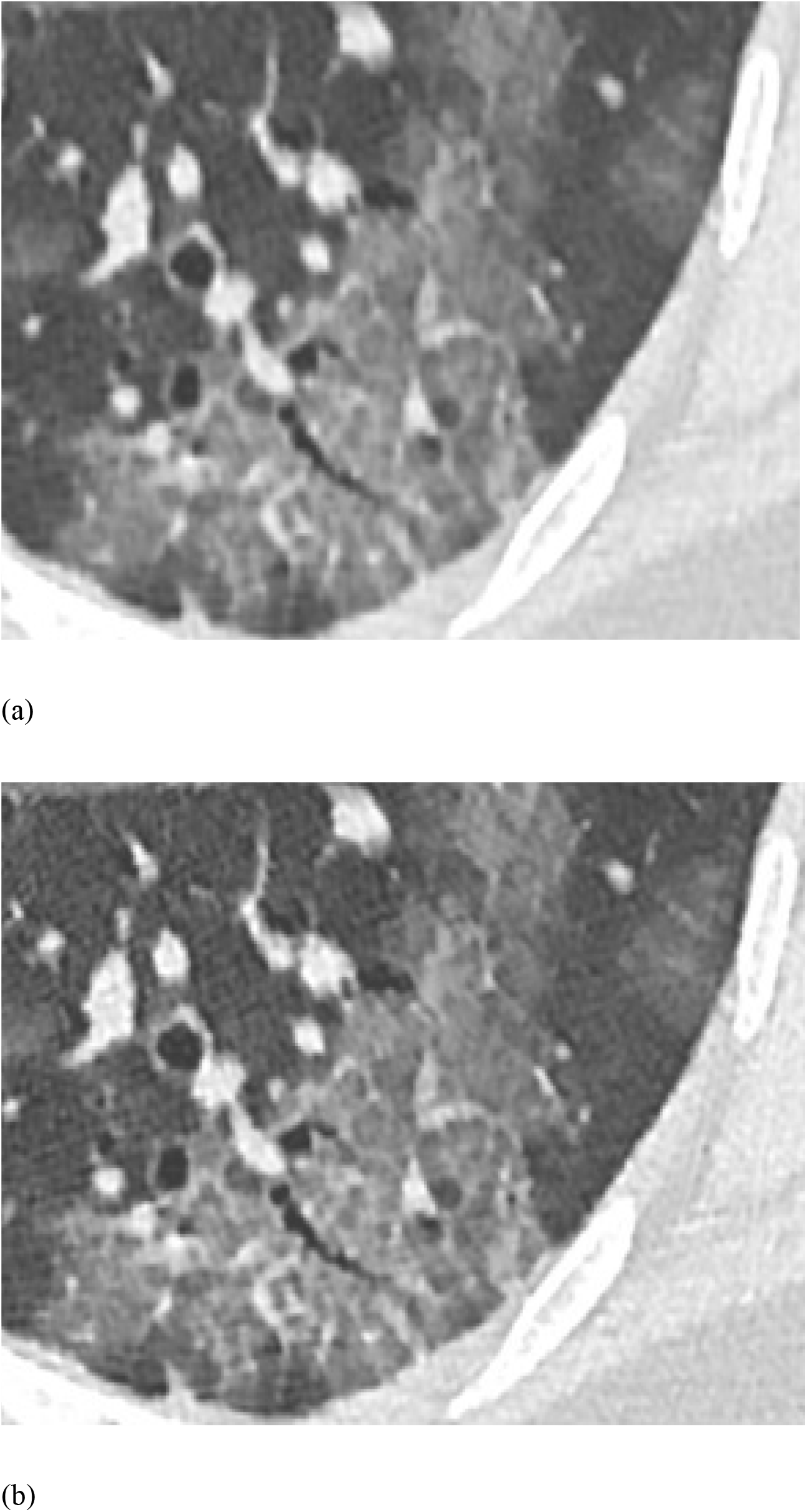
(a) C-HRCT and (b) U-HRCT images of a male 38-year-old patient with COVID-19. Crazy-paving patterns, interlobular septal thickening, air bronchograms, and smooth bronchial margins are clearly displayed onU-HRCT image

**Fig. 4.**
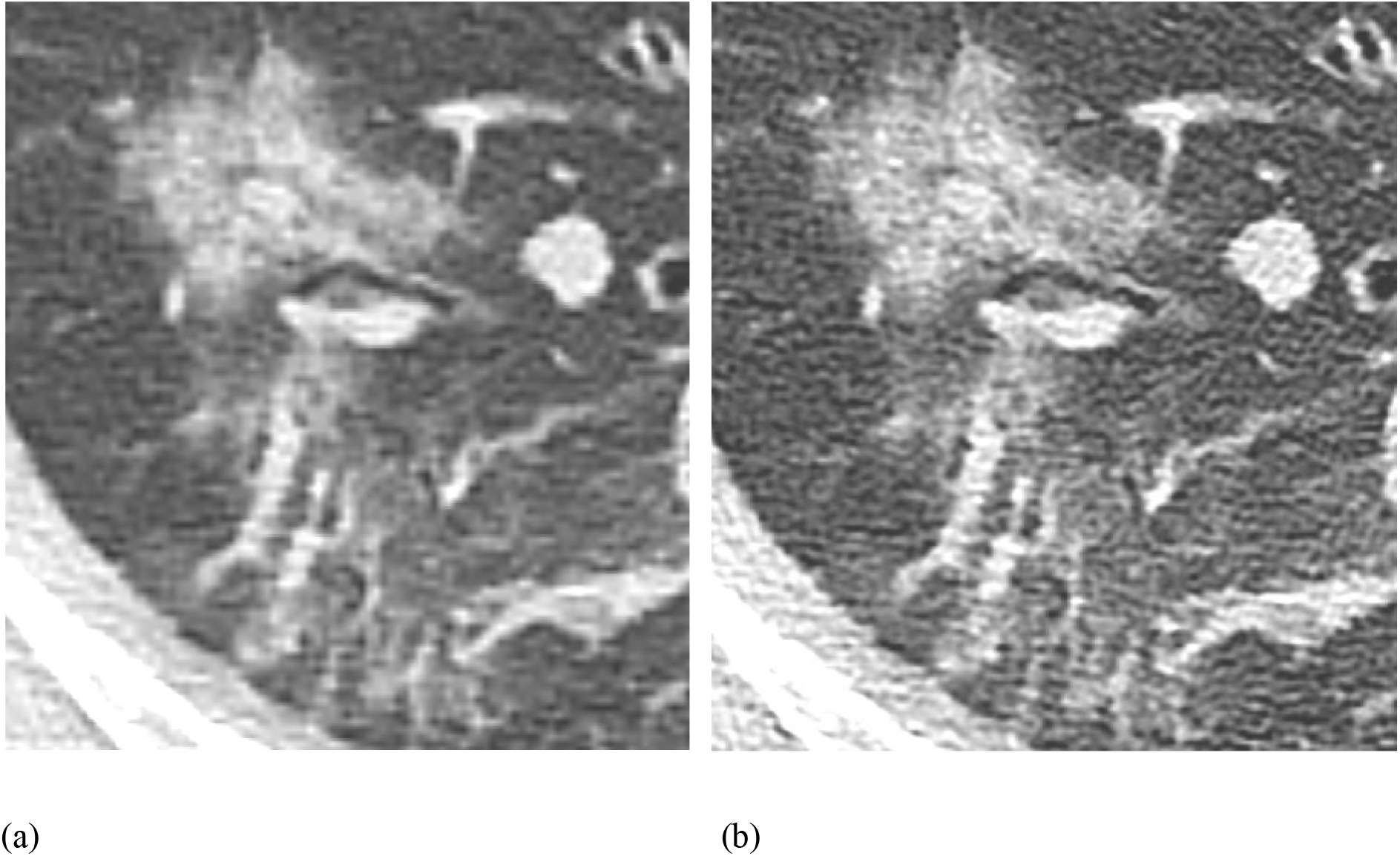
C-HRCT and (b) U-HRCT images of a female 55-year-old patient with COVID-19. Air bronchograms, smooth bronchial marginsin the lesion are clearly displayed in U-HRCT

There is no statistically significant difference between theC-HRCT and U-HRCT groups with respect to displaying crazy-paving patterns. There may be subjective factors involved in the interpretation of signs such as interlobular septal thickening and crazy-paving patterns.

This study had a few limitations. The number of patient samples included was small; consequently, statistical analysis may be biased. Retrospective target reconstruction was used in all cases, and no comparison with large-matrix U-HRCT target scan images was performed. Body mass index and radiation dose were not considered while evaluating image quality.

## Conclusions

U-HRCT image could reduce noise and improve SNR with an increase of the iDose^4^level. The iDose^4^-5 level had the highest U-HRCT score for c1linical applications. Peripleural distribution, thickening of blood vessels and interlobular septum, and crazy-paving pattern on U-HRCT are favorable signs for COVID-19. U-HRCT is superior to C-HRCT in displaying the blood vessels, bronchial walls, and interlobular septum for evaluating COVID-19.

## Data Availability

The datasets analyzed during the current study are fully available from the corresponding author on reasonable request.

## Declarations

### Ethics approval and consent to participate

The retrospective study was approved by the Institutional Review Board of Xiamen Universityand patients’ written consent to participate or parents/guardians (for anyone under the age of 16) was also obtained.

### Consent to publish

Not applicable.

### Availability of data and materials

The datasets analyzed during the current study are fullyavailable from the corresponding author on reasonable request. We confirmed that these patients have not yet been reported in any other submission by the authors.

### Authors’ Contributions

All authors have made substantial contributions to all of the following: (1) the conception and design of the study (LSM, LY, KJH and ZWG), or acquisition of data (LSM andLY), or analysis and interpretation of data (DSY, WJA and LSM), (2) drafting the article (LSM andLY) or revising it. critically for important intellectual content (ZWG and WJA), (3) final approval of the version to be submitted (LSM, LY, KJH, DSY, ZWG and WJA). Furthermore, each of the authors has read and concurs with the content in the manuscript.

## Acknowledgements

Not applicable.

## Abbreviations

COVID-19: Corona-virus disease 2019
SARS-CoV-2: Severe acute respiratory syndrome coronavirus 2
HRCT: High resolution computed tomography
C-HRCT: Conventional high resolution computed tomography
U-HRCT: Ultra high resolution computed tomography
FOV: Field of view
ROI: Region of interest
SD: Standard deviation
SNR: Signal-to-noise ratio
ROC: The receiver operating characteristic curve
AUC: Area under the curve

